# Genomic epidemiology links azole-resistant *Aspergillus fumigatus* hospital bioaerosols to chronic respiratory aspergillosis

**DOI:** 10.1101/2025.07.04.25330042

**Authors:** Amélie P Brackin, Rodrigo Leitao, Johanna Rhodes, Zain Chaudhry, David Connell, Samuel Hemmings, Jennifer M G Shelton, Matthew C Fisher, Darius Armstrong-James, Anand Shah

## Abstract

Drug-resistant infections caused by spores of the mould *Aspergillus fumigatus* pose a major challenge in managing chronic respiratory disease. Evidence shows that a substantial burden of aspergillosis is caused by strains that have evolved resistance to azole antifungal chemicals in the environment, however the contribution of local exposures to the colonisation of patients remains unclear. To investigate routes of acquisition, we whole-genome sequenced *A. fumigatus* isolates from individuals with chronic pulmonary fungal disease (*n*=182, 15 individuals), their homes (*n*=101, 10 homes), and hospital environments (*n*=102). These data were then integrated with retrospective sequence datasets enabling phylogenetic resolution across 912 genomes of UK *A. fumigatus*. We found high genetic diversity in clinical isolates, frequent mixed colonisation, and azole resistance in ∼25% of infections, particularly in those patients with cystic fibrosis (CF) and chronic pulmonary aspergillosis (CPA). The TR_34_/L98H *cyp51A* resistance allele, a well characterised marker of environmental adaptation to azole agricultural fungicides, was present in 25% of clinical azole-resistant strains. While azole-resistant *A. fumigatus* was detected in 6/10 homes, phylogenomic analysis revealed no clear genetic link between the home environment and clinical fungal lung isolates. *A. fumigatus* was prevalent in hospital environments, with azole-resistant isolates comprising 4.5% (*n*=9/202) of air and 3.4% (*n*=6/178) of soil isolates, predominantly harbouring the TR_34_/L98H allele. In contrast to homes, phylogenomic and pairwise SNP analysis revealed numerous clinical isolates with >97% genetic identity when compared to those isolated from the hospital environment and randomly chosen pairs of UK isolates. These findings indicate widespread exposure and potential nosocomial acquisition of drug-resistant genotypes of *A. fumigatus*, supporting the need for targeted environmental surveillance and mitigation of exposures in healthcare settings.

## Introduction

On October 25th, 2022, the World Health Organization (WHO) released its inaugural fungal priority pathogens list, calling for advanced diagnostics, robust surveillance, and accelerated antifungal development to address the escalating threat of drug resistance^1^. *Aspergillus fumigatus*, designated as one of four critical fungal pathogens of clinical priority, constitutes around 50% of the worldwide fungal disease burden, with high mortality rates and escalating drug resistance that disproportionately affects individuals with chronic respiratory diseases and viral pneumonias ^2^.

Triazole antifungal drugs are considered first-line therapy for aspergillosis, valued for their broad-spectrum antifungal activity, low toxicity, and oral availability, which has led to widespread clinical use^3–6^. However, the emergence of azole-resistant *A. fumigatus* (AR*Af*), characterised by polymorphisms in the *cyp51A* drug target, has significantly compromised treatment efficacy and worsened clinical outcomes. The widespread application of azoles in agriculture, horticulture, and various industrial biocides, including paints, coatings, wallpaper adhesives, textiles, and wood preservatives, has widely contaminated natural reservoirs ^7^, accelerating the dissemination of resistance traits and further complicating patient management. Chronic respiratory diseases complicated by fungal infection carry a heightened risk of AR*Af* acquisition as host defences often fail to fully eradicate the pathogen, frequently necessitating prolonged azole therapy. The resulting selective pressures, within a complex mosaic of physiological microenvironments shaped by spatial heterogeneity in immune responses, pharmacokinetics, and pharmacodynamics^8–10^, contribute to genetic diversity within the lung by driving evolution and persistence of azole-resistant populations ^11^. In London respiratory disease cohorts, AR*Af* has been detected in approximately 13% of individuals with aspergillosis, with over 40% of cystic fibrosis (CF) resistant cases attributed to the environmentally derived TR_34_/L98H genotype^12^, reflecting the substantial influence of environmental transmission on resistance patterns^13–15^. Genomic evidence of shared clonal backgrounds between clinical and environmental AR*Af* isolates^16,17^ implicates environmental transmission as a key pathway for resistance acquisition. UK-wide surveillance identified AR*Af* in ∼4% of air and ∼14% of soil samples, with minimal genomic differentiation between environmental and clinical strains, suggesting that over 40% of human AR*Af* infections may originate from environmental sources^18^. However, specific reservoirs and routes of acquisition remain incompletely defined, requiring further study to better understand the drivers of AR*Af* acquisition.

The TR_34_/L98H polymorphism has been detected in indoor environments, including soil and air from homes^19,20^ and healthcare settings^21–23^ positioning these spaces as significant sources of human exposure to AR*Af*. Multiple factors likely influence the introduction and persistence of AR*Af* indoors, such as building characteristics, occupant behaviours, air conditioning systems and outdoor spore infiltration^24,25^. Residual azoles present in commonly consumed foods, including teas, spices, and fruits^26–28^ may carry AR*Af* spores into indoor spaces, while damp, water-damaged areas create niches for fungal proliferation on substrates like wallpaper and paint, often treated with azole-based biocides ^29^. Routine domestic activities, such as vacuuming, dusting, and potting plants, further disperse spores, exacerbating exposure to *A. fumigatus* within indoor environments ^30,31^.

Environmental persistence may be exacerbated by host-derived transmission, where resistant inocula are released into the surroundings through coughing or exhalation^32^. Although in-host adaptation by *A. fumigatus* often results in traits such as reduced sporulation and slower growth^33,34^ which can impact fitness outside the host, compensatory mutations can mitigate these effects. Such mechanisms may sustain a cycle of azole resistance transmission and acquisition in high-exposure settings, creating distinct reservoirs of AR*Af*. These pathways are comparable to those observed in multidrug-resistant respiratory bacterial pathogens such as *Pseudomonas aeruginosa* and *Staphylococcus aureus*, which are known to contaminate healthcare environments *via* aerosolisation and contact^35–37^.

Currently, mycological surveillance in healthcare settings is limited by the high costs of implementation, the absence of standardised sampling protocols, and an emphasis on monitoring high-risk patient groups and a focus on specific, largely bacterial, pathogens ^38,39^. Routine screening is typically restricted to environments such as intensive care units, haemato-oncology, and burns wards, with fungal surveillance often limited to outbreak investigations or targeted research studies. This narrow approach may fail to detect clinically relevant fungal exposures among non-target patient populations, including those with chronic respiratory diseases, where risk of acquisition remains substantial.

In order to better understand acquisition of drug-resistant aspergillosis, we conducted a two-year ‘real-time’ epidemiological surveillance of *A. fumigatus* exposures by measuring the prevalence and type of azole resistance in individuals with chronic respiratory diseases and their close environments. High-resolution whole-genome sequencing (WGS) was conducted on clinical isolates from participants and combined with environmental WGS data from their homes and the hospital they attended to investigate possible ‘hot-spots’ of resistance in high-exposure settings. Comparator isolates from a UK-wide clinical and environmental *A. fumigatus* dataset of 616 genomes were included to contextualise phylogenomic diversity. This genomics-based approach enabled differentiation between clinical and environmental isolates with very high power, providing nuanced insights to inform targeted interventions for enhanced public health control measures.

## Results

### Clinical sample collection and characterisation of AR*Af*

Between September 2020 and April 2022, 296 remote (home) sputum sampling kits were distributed monthly to 55 individuals with chronic respiratory fungal disease attending a UK specialist tertiary respiratory centre for molecular epidemiology surveillance (Figure 1a). The cohort included individuals with cystic fibrosis (CF) (*n* = 9, 16.4%), chronic pulmonary aspergillosis (CPA) (*n* = 21, 38.2%), allergic bronchopulmonary aspergillosis (ABPA) (*n* = 23, 41.8%), *A. fumigatus* colonisation (*n* = 1, 1.8%), and chronic obstructive pulmonary disease (COPD) (*n* = 1, 1.8%) (Supplementary Table 1). Of the 296 kits distributed, 235 sputum samples were returned (achieving a 79% response rate), with *A. fumigatus* cultured from 27 participants (49%). Susceptibility testing of 321 *A. fumigatus* isolates against itraconazole (4 mg/L), voriconazole (2 mg/L), and posaconazole (0.5 mg/L) identified AR*Af* in 14 participants, corresponding to an overall prevalence of 25.5% (14/55) in the study cohort (Figure 1b). AR*Af* prevalence was highest in the 40–59 age group; CPA participants had the highest resistance rate (42.9%), followed by CF (33.3%) and ABPA (4.3%). Multivariate logistic regression (Supplementary Figure 1) confirmed CPA and CF as strong independent predictors of resistance.

**Figure 1.**
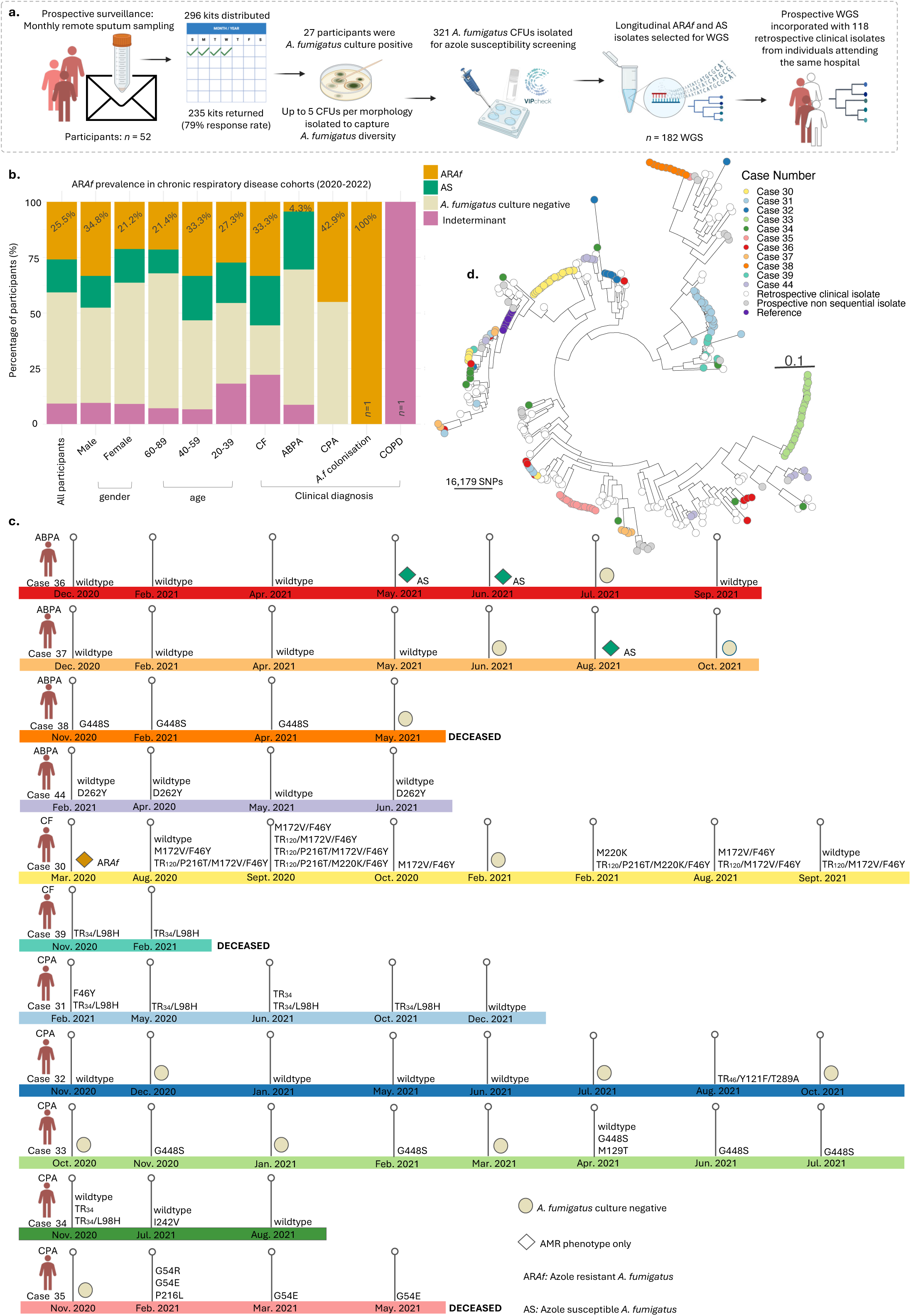
Prospective surveillance of azole resistance in individuals with chronic respiratory diseases. (a) Schematic representation of the study design and experimental framework. Workflow included sample kit preparation and delivery, sputum sample processing and fungal culture, drug susceptibility testing and DNA extraction for WGS analysis. See methods section for detailed information. (b) Prevalence of azole resistance among participants in the respiratory disease cohort who had at least one azole-resistant isolate recovered from longitudinal samples. Resistance prevalence is further categorised by gender, age, and clinical diagnosis. (c) Distribution of *cyp51a* polymorphisms detected during prospective surveillance. (d) Phylogenetic tree illustrating the evolutionary relationships of clinical *A. fumigatus* isolates. Coloured circles on the branches represent the Case ID for prospective study participants. To strengthen phylogenetic analysis, genome sequences were integrated into an existing WGS data repository, which included 118 clinical genomes (white circles) from individuals with chronic respiratory diseases attending the same hospital.

A subset of 182 clinical isolates from 15 individuals, each with at least two sequential isolates sampled 1–8 months apart, underwent WGS (Supplementary Table 2). Sixteen distinct *cyp51A* genotypes were identified, indicating within-cohort genetic diversity (Figure 1c). Five individuals (Cases 34, 39, 41, 42, 43) harboured isolates distributed across multiple phylogenetic clusters, consistent with intrahost genetic diversity (Figure 1d). In contrast, 10 individuals harboured sequential isolates that clustered phylogenetically, suggesting possible strain stability and persistence. The TR_120_ polymorphism, recently reported in Dutch isolates ^40^, was identified in a UK CF patient, marking its first documented case in the UK. These isolates, collected over 11 months, exhibited distinct SNP profiles and resistance phenotypes yet clustered phylogenetically. Inclusion of retrospective genomes identified a TR_120_/M172/F46Y isolate from Case 30, collected in 2016, clustering with both prospective TR_120_/M172/F46Y isolates and TR_120_ isolates with or without additional polymorphisms, indicating long-term persistence and micro-evolution of the variant. Similarly, a retrospective TR_34_/L98H isolate from Case 31 clustered with prospective azole-resistant isolates, indicating likely intra-host persistence. Case 31 also harboured a phylogenetically distinct TR_34_ variant without L98H, highlighting the coexistence of multiple genetically distinct strains within the host. Among the 15 individuals, the TR_34_/L98H genotype was identified in 26.6% (*n* = 4) of cases, with all isolates resistant to at least one clinically-used azole. An isolate carrying a TR_46_/Y121F/T289A genotype isolate was recovered from Case 32, marking the first report of this genotype in London cohorts.

### Home environmental surveillance and characterisation of AR*Af*

To understand whether home environments were important in AR*Af* acquisition, targeted home environmental surveillance was performed in specific participants isolating AR*Af* and non-AR*Af* clinical *A. fumigatus* isolates to enable genome sequencing for molecular epidemiology. A schematic of the experimental workflow is shown in Figure 2a. *A. fumigatus* bioburden varied across homes (Figure 2b), with no significant association between environmental and sputum CFU counts (Spearman’s ρ = 0.267, p = 0.455). Of 438 *A. fumigatus* isolates, AR*Af* was detected in 6 of 10 homes, representing 3.9% (17/438) overall: 2.7% (12/438) in soil, 0.7% (3/438) in air, and 0.5% (2/438) in dust. WGS of 101 isolates, selected to represent both wildtype and azole-resistant *A. fumigatus* in the home environment, revealed eight *cyp51A* genotypes (Supplementary Table 3). Multivariate Discriminant Analysis of Principal Components (DAPC) was performed to evaluate household-specific adaptation or transmission, but no clustering was observed (Figure 2c). Phylogenetic analysis (median bootstrap support 98%) showed that home isolates were distributed across multiple branches, consistent with extensive genetic diversity and a cosmopolitan population structure. Inclusion of participants’ clinical isolates in the phylogenetic analysis revealed no close genetic relationships between clinical and home-derived isolates (Figure 2d).

**Figure 2.**
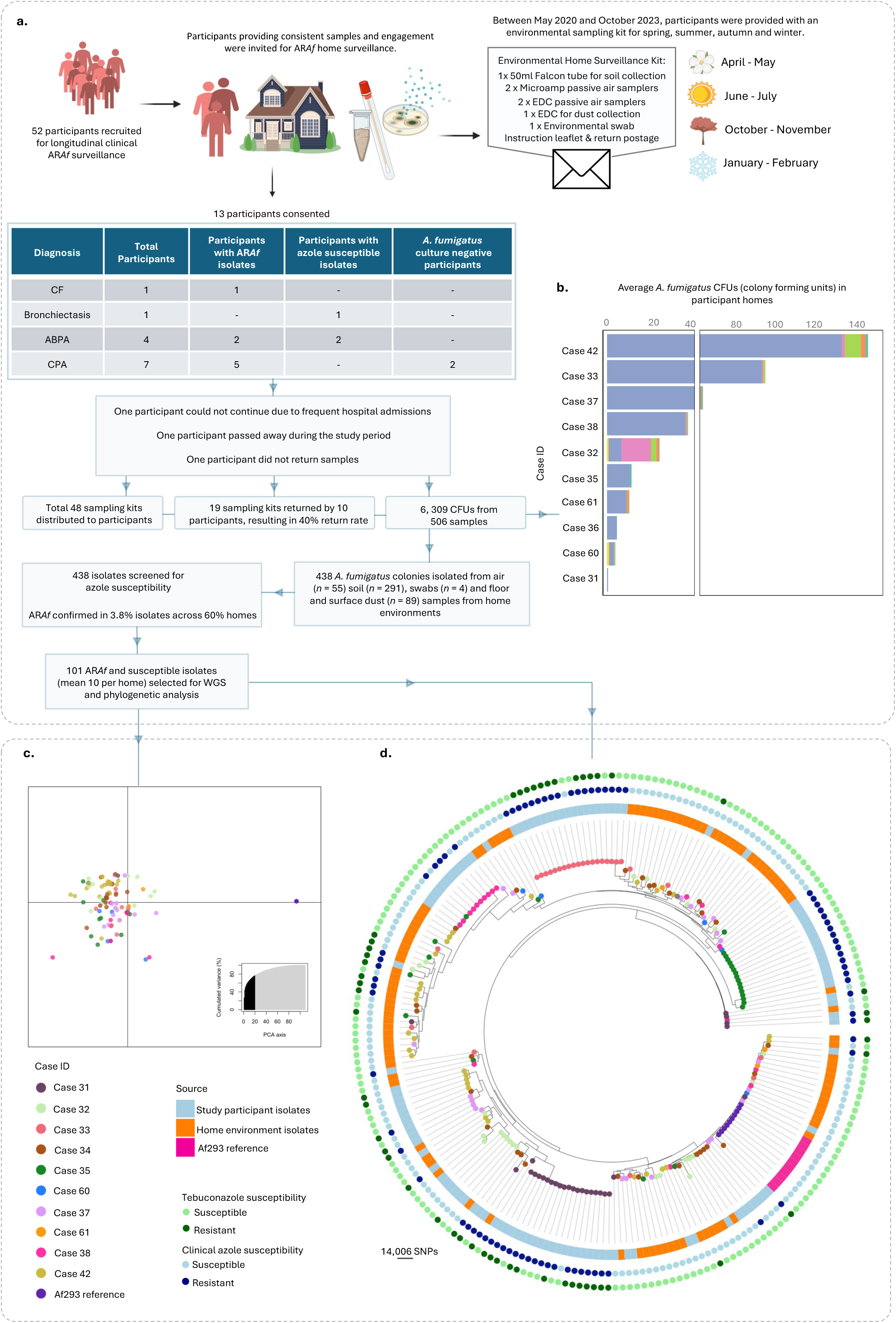
Environmental surveillance of azole resistance in study participants homes. (a) Schematic representation of the study design and experimental framework. Workflow included sample kit preparation and delivery, environmental sample processing and fungal culture, drug susceptibility testing and DNA extraction for WGS analysis. See methods section for detailed information. (b) Average CFU counts across participating home environments. (c) DAPC based on SNP variants from of *A. fumigatus* isolates collected from air, soil and dust of participant homes. Coloured points represent the homes of each Case ID. The Af293 refence genomes (*n*=10) are shown as purple points. The inset shows the cumulative variance explained by the principal component axes, indicating the proportion of total variance captured by the analysis. (d) Phylogenetic tree showing evolutionary relationships between *A. fumigatus* isolates from participant homes and corresponding clinical samples. Coloured circles on the branches represent the source of the isolate (Case ID). The inner circle indicates the source, either clinical or home environment. The outer rings indicate susceptibility profiles for tebuconazole (green) and the clinical azoles (blue), with darker colours representing resistance.

### Hospital environment surveillance and characterisation of AR*Af*

We subsequently performed environmental surveillance across multiple sites at a London hospital attended by study participants (Figure 3) in order to analyse azole-resistance incidence within healthcare settings and their potential importance in the acquisition of AR*Af*. A schematic of the experimental workflow is shown in Figure 4a. *A. fumigatus* bioburden varied across sites (Figure 4b), with the highest airborne concentrations observed in a reception area (median 6 CFUs/m³), followed by basement areas, staff offices (both with median 3 CFUs/m³), and outpatient departments (median 2.5 CFUs/m³). Indoor flower beds had a higher proportion of *A. fumigatus* positive samples, with those in the reception area showing the highest concentration (median 3.5 CFUs/g; range 0–75 CFUs/g). Tebuconazole-resistance was analysed using the TebuCheck protocol to screen for AR*Af* ^41^ with detection in 4.5% (*n*=9/202) of air isolates and 3.4% (*n*=6/178) of soil isolates. All clinical azole-resistant isolates also exhibited cross-resistance to tebuconazole (6 mg/L). Resistant isolates were spatiotemporally distributed, predominantly in the hospital basement, outpatient department and reception area.

**Figure 3.**
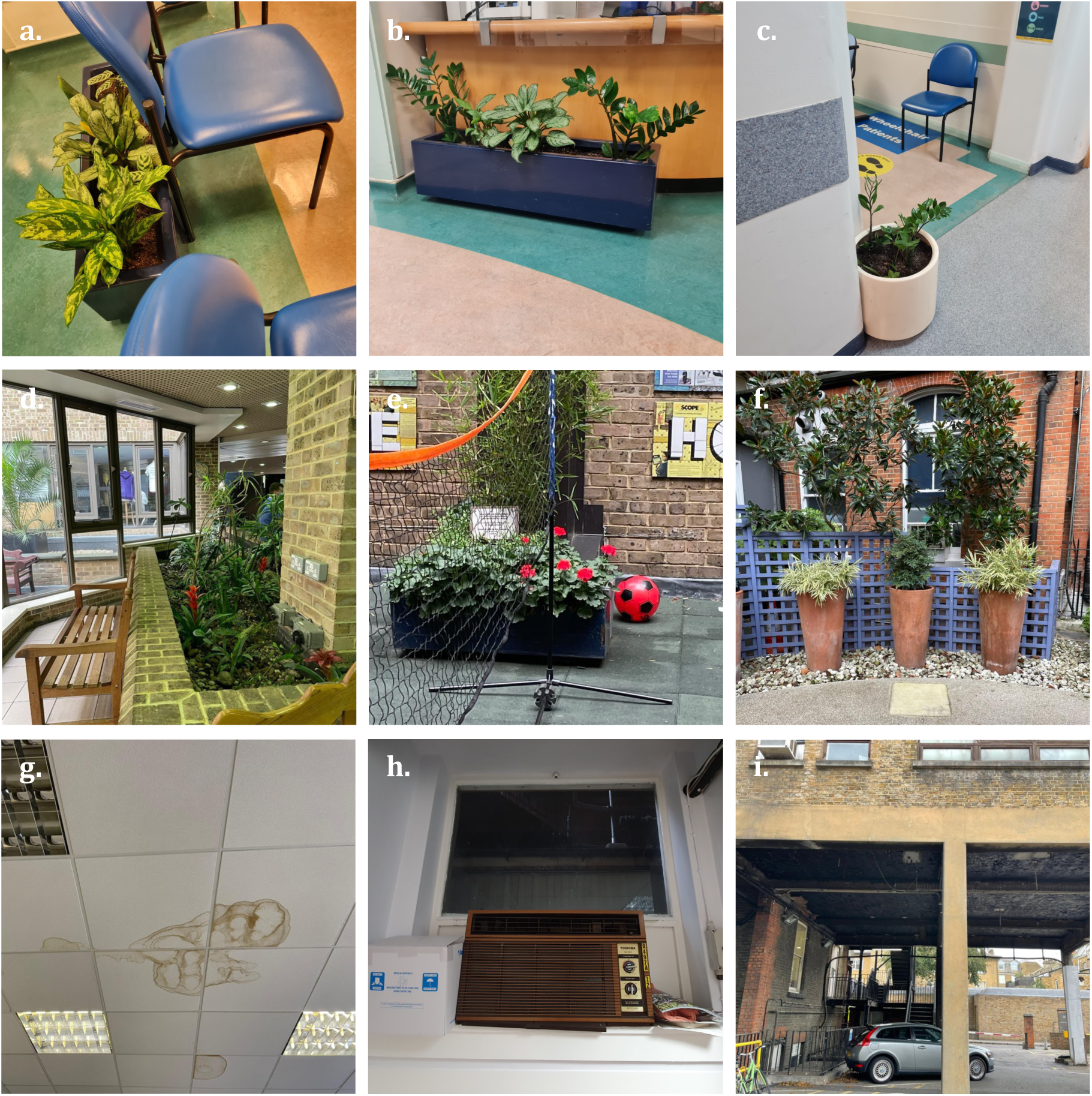
Hospital sampling locations. Soil and air samples were collected from multiple locations within the hospital to assess the presence and distribution of *A. fumigatus*. Soil samples were collected from indoor and outdoor potted plants and garden spaces, including patient waiting areas (a-c), a reception area (d) and outdoor courtyards (e-f). Air samples were also collected from locations with potential air quality issues, including areas with signs of dampness or old air conditioning units: ceiling with water damage (g) and a window-mounted old air conditioning unit (h). External control located at the back of the hospital in a staff car park (i).

**Figure 4.**
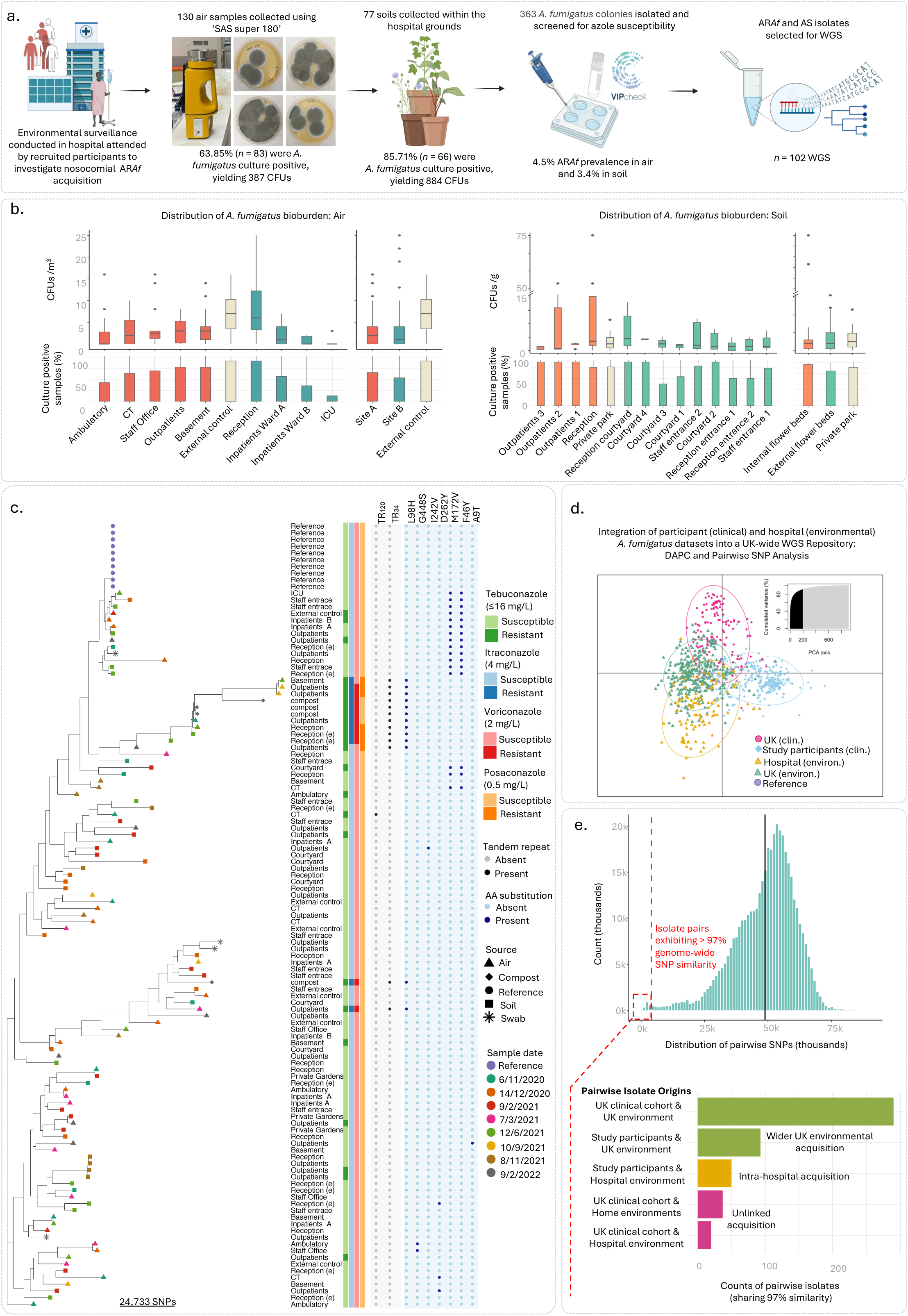
Environmental surveillance of azole resistance in a central London Hospital. (a) Schematic overview of the study design and experimental workflow, including environmental sampling, fungal culture, drug susceptibility testing, and DNA extraction for whole-genome sequencing. Full methodological details are provided in the Methods section. (b) Spatial distribution of *A. fumigatus* in hospital air and soil samples. Left: percentage of culture-positive air samples collected from two hospital sites, the Victorian-era Site A (red) and the modern Site B (blue), with an external control site (beige). Colony forming units (CFUs/m³) in air samples from the same locations, shown as box plots indicating the median, interquartile range, and outliers. Right: percentage of culture-positive soil samples collected from internal (orange) and external (green) flower beds, with a private London park included as an external control (beige). Colony-forming units (CFUs/g) for soil samples are shown as box plots. (c) Phylogenetic tree showing evolutionary relationships among hospital environment *A. fumigatus* isolates. *Cyp51A* alleles are marked by grey points (tandem repeats) and blue points (amino acid substitutions). (d) Discriminant analysis of principal components (DAPC) plot showing genetic clustering of hospital environmental isolates, study participant isolates and UK-wide environmental and clinical isolates. (e) Counts of pairwise comparisons with >97% genomic similarity, including their source.

WGS of a representative set of 102 hospital-derived environmental isolates was performed to investigate the genetic background of AR*Af* and to identify potential nosocomial acquisition. Genotypes and their resistance profiles are shown in Supplementary Table 4. Pairwise SNP analysis among hospital environmental isolates showed an average divergence of 47,971 SNPs, which is similar to that found more widely in the UK (average divergence 48,838 SNPs). Ten isolate pairs met the genetic similarity threshold (<2,577 SNPs, >97% of maximum observed diversity). Four pairs originated from the same sample, likely due to replication during culture, while the remaining six pairs were recovered from different locations or time points, indicating the widespread circulation of highly-related isolates. The TR_34_/L98H polymorphism was identified in 8/102 (8%) isolates, primarily from air samples collected in the basement, outpatients department, and reception, all of which clustered phylogenetically (Figure 4c), making this the second most common genetic polymorphism after M172V/F46Y. Six of the TR_34_/L98H isolates were resistant to at least one clinical azole and tebuconazole (up to 16 mg/L). Azole-susceptible isolate pairs harbouring the G448S mutation (exhibiting >94% genetic similarity), showed a comparable spatiotemporal distribution and were recovered from air samples in staff offices and outpatient areas in December 2020 and March 2021, respectively.

### Incorporation of 616 UK comparator whole-genome sequences

To achieve greater longitudinal resolution of intra-host AR*Af* dynamics, an additional 118 retrospective clinical genomes from respiratory disease cohorts at the same central London hospital were incorporated, expanding the clinical dataset to 300 genomes from 83 individuals collected between 2015–2022. To further enhance phylogenetic resolution and investigate environmental AR*Af* transmission, 151 UK-wide clinical isolates and 347 environmental isolates were included as comparators. DAPC analysis was used to illustrate the genetic relationships between the clinical and environmental clusters (Figure 4d). Study participants’ isolates overlapped with UK clinical and environmental isolates, indicating acquisition from multiple sources. Hospital environmental isolates clustered separately but showed partial overlap with study participants’ isolates, supporting the role of hospital environments as potential reservoirs for AR*Af* exposure and acquisition. This observation was in sharp contrast with participants’ homes, where no closely-related isolates were identified (Figure 2c). Pairwise genetic similarity (>97% identity, <1,474 unique SNPs) identified 994 closely-related isolate pairs in the UK dataset. Analysis of these pairs aimed to resolve the most likely environmental source of clinical *A. fumigatus* isolates. Most clinical pairs across the wider UK dataset (*n* = 290/994) showed close relationships to isolates from the wider environment, indicating this as the predominant reservoir of exposure (Figure 4e). However, 49 pairs showed >97% identity between clinical isolates from this study’s participants, and their London hospital environment, suggesting possible intra-hospital transmission. Of these, 28 comparisons showed clinical TR_34_/L98H, G448S, and D262Y that were also identified in the hospital environment, with >97% identity amongst clinical and environmental isolates. Strikingly, within the Central London Hospital cohort, 41% (*n* = 7/17) of individuals carried isolates with >97% genetic identity that were exclusive to the hospital environment, while 35% (*n* = 6) had isolates linked to the wider UK environment but not their homes.

Notably, phylogenomic analysis of the complete UK dataset, comprising 912 isolates, revealed tightly clustered clinical and hospital environmental isolates from this study, indicative of nosocomial acquisition of infection (Figure 5a). The UK phylogeny, accessible *via* a Microreact project (https://microreact.org/project/ohanRa45Se2kRQQugq6mK9-uk), showed a median bootstrap support of 78% across the tree, with nosocomial clusters supported by 100% bootstrap values (Supplementary Figure 2).

**Figure 5.**
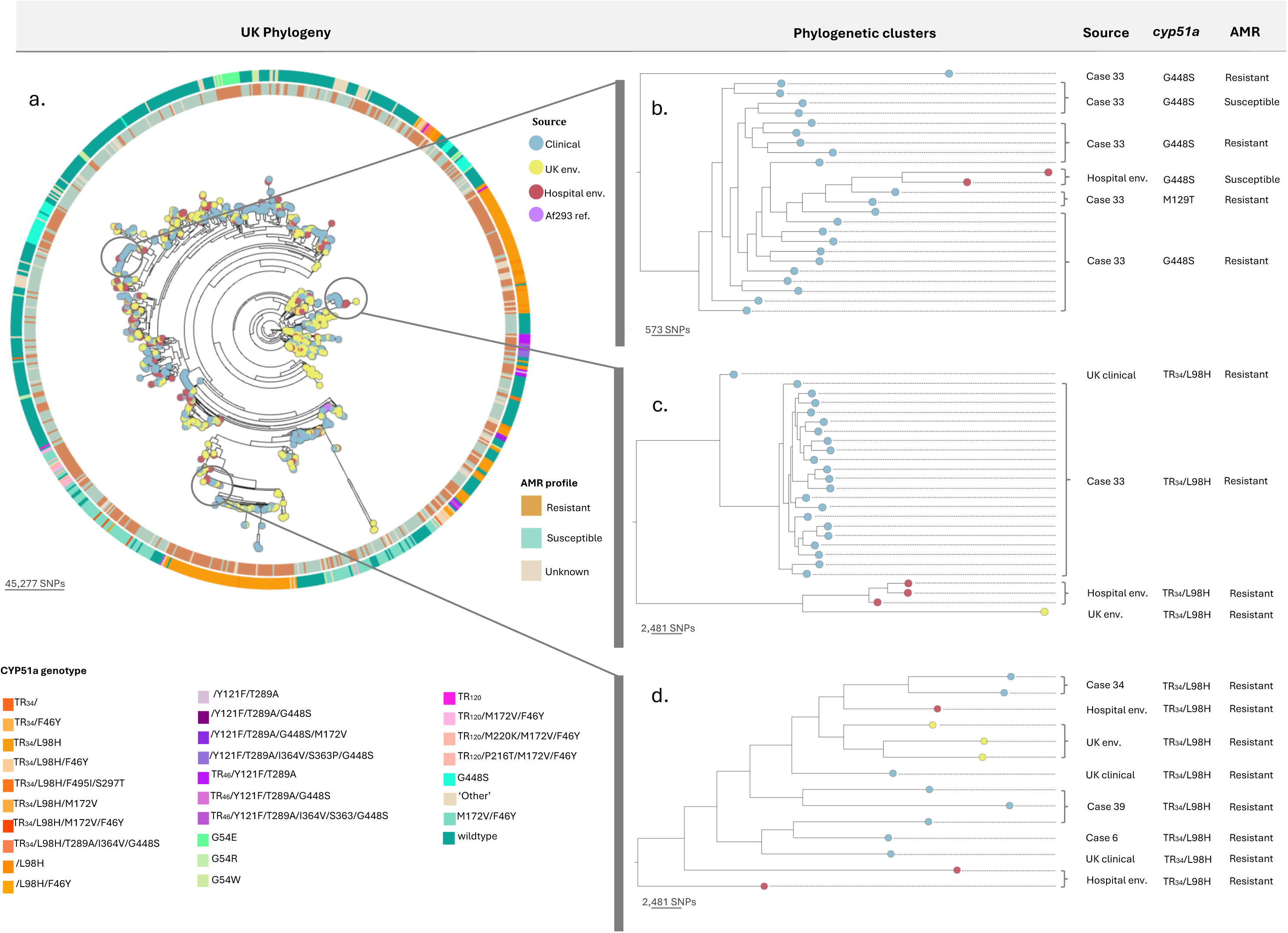
Phylogenetic analysis and genotypic characterisation of UK *A. fumigatus* clinical and environmental isolates. Phylogenetic relationships among UK clinical and environmental isolates (a) with highlighted phylogenetic clusters showing epidemiological links between clinical and hospital environmental *A. fumigatus* (b-d).

Case 33 (a CPA participant) harboured AR*Af* isolates with the cyp51A G448S allele over ten months (October 2020 – July 2021), showing an average pairwise divergence of 2,438 SNPs (95% identity) and phylogenetic clustering across this time-series (Figure 5b). The G448S allele is normally associated with in-host evolution^42^ yet occurred in hospital air samples collected in December 2020 and March 2021. These G448S-bearing isolates from the hospital environment were shown to phylogenetically cluster with Case 33 isolates, but were separated by an average of 4,199 SNPs (91.4% genetic similarity; Figure 5b). This phylogenetic clustering indicates possible hospital acquisition; however, the genetic distance suggests the absence of recent demonstrable direct transmission. Other clusters of hospital clinical and environmental TR_34_/L98H isolates similarly point to potential hospital acquisition (Figures 5c-d). However, the presence of closely-related isolates from other UK clinical and environmental sources indicates broader occurrence of these genotypes and acquisition from exposures outside the hospital cannot be ruled out.

## Discussion

Azole-resistant *A. fumigatus* is an increasing global health threat, particularly complicating chronic *Aspergillus-*related lung disease and resulting in significant morbidity and mortality. In this study, using real-time microbiological surveillance integrated with WGS, we identify healthcare environments as reservoirs of AR*Af* bioaerosol exposures. That several clinical isolates shared >95% genetic similarity to hospital-derived environmental isolates is consistent with nosocomial transmission pathways in settings where there is a concentration of at-risk individuals. Environmental nosocomial surveillance in high-traffic areas performed in our study demonstrated the pervasive presence of *A. fumigatus* and AR*Af* within healthcare settings, including those areas of the hospital with stringent protective measures ^43–47^ (but not intensive care settings). Surveillance of the hospital environment revealed genetically-similar AR*Af* across multiple locations and time points, demonstrating that AR*Af* strains can survive long-term in hospital settings. These findings clearly highlight the need for robust surveillance in order to determine the extent that such exposures occur in this, and other, healthcare settings and, where observed, what strategies to mitigate exposures in vulnerable patient populations can be used.

Triazole-resistant isolates in the hospital setting were found at prevalences that broadly matched those reported in the wider UK environment of 3–6% ^18,48–50^ confirming that AR*Af* strains were well-established in particular healthcare environments – the hospital basement, outpatient department and reception area - highlighting the potential for localised transmission and an ongoing risk of repeated patient exposure to AR*Af*. Patients with lung disease and aspergillosis may further transmit strains into the hospital environment through coughing and expectoration^32^. If patients harbour isolates that are drug-resistant (either from *in-host* evolution or environment acquisition), these inocula may contribute to the establishment of distinct healthcare-specific environmental reservoirs of azole-resistance and a bidirectional cycle of contamination and acquisition.

High genetic similarity between clinical and hospital environmental isolates characterised by TR_34_/L98H and G448S suggest interlinked transmission pathways facilitated through direct or indirect routes of exposure. Although G448S is predominantly linked to *in-host* evolution, independent studies documenting *cyp51a* point mutations in environmental samples suggest that such mutations can arise independently of clinical pressures, complicating the distinction between patient-derived and environmental resistance pathways ^51–55^. This is further supported by our detection of the first UK hospital environmental isolate carrying the TR_120_ allele, previously documented only in Dutch CF isolates and associated with *de novo* evolution during prolonged azole therapy^40^. In Case 30, phylogenetic analysis of isolates collected over 11 months showed high relatedness (100% bootstrap support), with variable non-synonymous SNP profiles both with and without TR_120_. Genomic analysis confirmed that clinical TR_120_ isolates were genetically distinct from the hospital environment-derived TR_120_ isolate, suggesting either patient-to-environment transmission followed by compensatory adaptation, independent emergence under hospital selective pressures (challenging the assumption that TR_120_ mutations arise exclusively within the host), or the existence of unsampled intermediary isolates linking these reservoirs. The absence of a reliable molecular clock for *A. fumigatus* limits precise reconstruction of transmission pathways.

While targeted interventions to mitigate exposure and acquisition of AR*Af* by at-risk individuals may be achievable in controlled hospital environments, high-traffic areas such as outpatient clinics and waiting areas present greater challenges for effective regulation. In this context, the integration of remote patient monitoring, including at-home testing, offers a practical strategy to reduce hospital visits, limit exposure in high-risk settings and disrupt potential nosocomial transmission networks. Although surveillance efforts often intensify during periods of hospital construction ^56^, the persistent presence of *Aspergillus* spores as shown in our study highlights the importance of continuous environmental monitoring. Long-term mitigation will require infrastructure modifications to minimise fungal persistence and a reassessment of environmental features such as indoor planting, even in areas not traditionally classified as high risk ^57^. Further mitigation strategies such as air filtration, ventilation and specific infection control protocols for individuals with known AR*Af* additionally require consideration in this context.

Our study further demonstrates the high incidence of AR*Af* in individuals with chronic respiratory fungal disease such as CPA with variable intra-host *A. fumigatus* phylogenetic diversity reflecting dynamic pathogen populations within the human host. Our study employed frequent high-volume culture sampling previously shown to improve microbiological *A. fumigatus* yield emphasising the need for comprehensive sampling to support robust epidemiological conclusions on resistance acquisition ^58,59^. Closely-related serial isolates in chronic respiratory aspergillosis suggest repeated environmental acquisition or (more likely) intra-host persistence under prolonged selective drug pressure. In Case 31, TR_34_/L98H isolates detected from 2016-2023 exhibited high phylogenetic relatedness (100% bootstrap support), with a TR_34_ isolate lacking L98H also identified, marking the first clinical report of this allele. High rates of recombination caused by the sexual cycle of *A. fumigatus*, primarily described in environmental settings ^60,61^, have been shown to generate resistance genotypes such as TR_34_/L98H from parental strains carrying separate TR_34_ and L98H alleles ^62–64^. The phylogenetic relationship between TR_34_ and TR_34_/L98H isolates raises the possibility of acquisition from a persistent environmental source or, alternatively, a recombination event occurring within the host. These findings demonstrate the adaptive versatility of AR*Af,* whether through environmental persistence, recombination, or repeated acquisition. The detection of TR_34_/L98H in hospital environments, despite no phylogenetic link to Case 31 clinical isolates, suggests episodic exposure may lead to recurrent acquisition and potential within-host adaptation.

Genomic analysis identified the TR_34_/L98H resistance mechanism in 25% of azole-resistant clinical cases and confirmed the second occurrence of the TR_46_/Y121F/T289A mutation within a UK clinical cohort, underscoring the contribution of environmental AR*Af* to resistance acquisition. The high prevalence of TR_34_/L98H in the chronic *Aspergillus*-related lung disease cohort reflects its widespread occurrence as a globally-dominant resistance mechanism^16,17,65,66–68^. Detection of the TR_46_/Y121F/T289A allele, a resistance mechanism with high-level voriconazole resistance in Case 32 signals its emergence in the UK, echoing reports from Europe^59,69–72^, Asia^73–75^ and Africa^76^. Although first identified in a clinical isolate in Manchester in 2017^77^, retrospective surveillance from 1998–2017 found no cases among 1,469 isolates in London cohorts^78^. The first environmental detection in 2019, involving six isolates from soil, compost, and hospital flower beds in London, confirmed local presence of TR_46_/Y121F/T289A^48^. By 2023, a national surveillance study found TR_46_/Y121F/T289A in 6% of aerosolised *A. fumigatus* isolates, compared to 59% for TR_34_/L98H^49^, suggesting a trajectory of broader dissemination similar to that previously observed for TR_34_/L98H. This is of significant concern for global use of voriconazole which is a first-line antifungal therapy for invasive aspergillosis.

In conclusion, using real-time epidemiological surveillance we show that hospital environments may serve as unique reservoirs of AR*Af*, maintained through the introduction of host-derived isolates and sustained local transmission cycles. The complex pathological landscape of chronic respiratory diseases, marked by fluctuating inflammation, microbial burden, and prolonged antifungal exposure, may further facilitate the persistence of genetically diverse populations and contribute to resistance evolution across both environmental and clinical reservoirs. The role of nosocomial AR*Af* reservoirs in sustaining resistant populations within healthcare environments where at-risk patients concentrate, underscores the need for expanded genomic surveillance across diverse settings and cohorts in order to better understand the mitigation strategies that are needed to protect at-risk patients.

## Methods

### Participant selection and inclusion criteria for prospective surveillance

Participants aged ≥16 years with a confirmed diagnosis of chronic *Aspergillus*-related lung disease on a background of CF, non-CF bronchiectasis, CPA, COPD or other chronic lung conditions were recruited within the domains of a central London Hospital. Informed consent was obtained before enrolment, and detailed clinical data were collected, including demographics, medical history, respiratory health, and antifungal treatment history. Sputum samples were collected at the initial screening visit and monthly *via* remote sampling.

### Sputum sample collection and processing

Monthly sputum samples were collected remotely from 55 participants diagnosed with chronic *Aspergillus*-related lung disease. Each participant received a home sampling kit containing a sterile 30 mL collection tube (Thermo Scientific™, Sterilin™), Category 2 UN73373 packaging, and a leaflet with collection and mailing instructions. Samples were homogenized in a 1:1 (v:v) ratio with Mucolyse Sputum Digestant (Pro-Lab Diagnostics, UK) and incubated at room temperature for 20 minutes. A 200 µL aliquot of digested sputum was inoculated onto Sabouraud dextrose agar (SDA) supplemented with 50 mg/L chloramphenicol (Oxoid, Basingstoke, UK) and incubated at 45°C to select for *A. fumigatus*.

### Home Surveillance: Participant selection criteria

Participants were contacted *via* phone or email to explain the study’s purpose and assess their interest. Thirteen individuals were selected for home environmental monitoring, prioritising those with ≥3 culture-positive samples to examine *A. fumigatus* diversity. This included eight participants with AR*Af* or confirmed *cyp51A* resistance alleles to investigate potential home transmission pathways. As controls, three participants with azole-susceptible *cyp51A* wildtype isolates and two with culture-negative samples were included to assess AR*Af* burden in non-resistant cases. Among the selected participants, diagnoses included CF (*n* = 1), CPA (*n* = 7), ABPA (*n* = 4), and bronchiectasis (*n* = 1). During the study, one participant withdrew due to regular inpatient stays, and one died.

### Home surveillance: Sample Collection

Surveillance kits were provided every three months to capture seasonal variation and included materials for collecting indoor air, soil, dust, and swab samples, along with pre-labelled packaging for traceability and a detailed instruction leaflet. Samples collected from January– February, April–May, June–July, and October–November were classified as winter, spring, summer, and autumn, respectively. Returned samples were processed immediately or stored at 4°C until analysis. To optimise *A. fumigatus* recovery, participants used two validated passive air samplers ^49,79,80^: MicroAmp™ clear adhesive film (Thermo Fisher Scientific, UK) and electrostatic dust collectors (EDCs) (Liefheit, UK). The EDCs were sterilised by autoclaving at 121°C for 15 minutes prior to distribution. Each kit contained two of each sampler, which were placed at ∼1.2 meters height to simulate breathing zone exposure. The adhesive film and EDCs were exposed to indoor air for 14 days before being sealed in sterile ziplock bags. Settled dust was collected using an EDC attached to a vacuum nozzle or by sweeping surfaces. Surface soil samples were collected from indoor potted plants or, if unavailable, from outdoor flower beds, using sterile 50 mL Falcon tubes. Kits included FFP2 NR face masks (Medisave, UK) and latex-free gloves for safety during collection.

### Home surveillance: Sample processing

*A. fumigatus* colonies were recovered from MicroAmp™ adhesive air samplers (Applied Biosystems, UK) using previously described protocols ^49^. EDC samples were washed in 50 mL of 0.85% NaCl with 0.5% Tween-20, shaken at 250 rpm for 60 minutes at room temperature, centrifuged, and the supernatant removed to concentrate particulate matter before resuspension. Aliquots (150 µL) were inoculated in triplicate onto culture plates. Surface swab samples were washed in 1 mL of 0.85% NaCl with 0.5% Tween-20, vortexed for 1 minute, and 200 µL aliquots were plated in triplicate. Soil samples (2 g) were suspended in 8 mL of 0.85% NaCl with 0.5% Tween-20, vortexed, and 200 µL of supernatant was plated in triplicate. All culture plates were incubated at 45°C and inspected daily for fungal growth.

### Hospital surveillance: Sampling locations

Seasonal air and soil samples were collected between November 2020 and February 2022 from a central London hospital comprising two sites: Site A, a Victorian-era building still in use for outpatient clinics, and Site B, a modern extension constructed in the early 1990s housing inpatient respiratory and cardiac wards. Air samples from Site A were collected from three outpatient clinics, a staff office, and the basement. Soil samples were obtained from internal flower beds adjacent to an outpatient clinic and external flower beds near the staff entrance. In Site B, air samples were collected from two inpatient wards, the Intensive Care Unit (ICU), and the hospital reception. Soil samples were taken from internal and external flower beds at the hospital reception and two external courtyards. Additional air samples were collected from an external staff car park.

### Hospital surveillance: Sample Collection and processing

Air samples were collected using a ‘SAS Super 180’ (Cherwell Laboratories, UK) air sampler positioned at approximately 1.2 meters height. Contact plates (90 mm) containing SDA supplemented with antibiotics (16 mg/L penicillin and 16 mg/L streptomycin) were placed on the aspirating head, sampling 1000 L of air over 5 minutes. Plates were sealed with Parafilm, stored in ziplock bags, and incubated at 45°C for 7 days, with daily inspections for colony growth. For soil sampling, 2 g of surface soil from internal and external hospital flower beds was suspended in 8 mL of sterile suspension buffer (0.85% NaCl, 0.01% Tween®-20, ThermoFisher Scientific) as previously described ^81^. Briefly, a 200 µL supernatant aliquot was plated onto SDA with antibiotics and incubated at 45°C for up to 7 days to isolate *A. fumigatus*.

### *A. fumigatus* isolate collection and azole susceptibility screening

Colony counts were recorded for clinical and environmental samples, with up to five colonies per morphology isolated onto SDA plates to capture diversity. Azole susceptibility profiles of 321 clinical and 826 environmental isolates were assessed using 4-well VIPCheck™ plates (Mediaproducts BV, Netherlands) containing agar supplemented with itraconazole (4 mg/L), voriconazole (2 mg/L), posaconazole (0.5 mg/L), and an azole-free control, following established protocols ^82^

### DNA extraction and WGS

Clinical and environmental isolates were selected to represent host-specific and spatiotemporal diversity, ensuring a 1:1 ratio of azole-susceptible to azole-resistant isolates per sample type. DNA was extracted using MasterPure Yeast Cell Lysis Solution (Epicentre Biotechnologies, UK) with bead-beating (1.0 mm zirconia beads, Thistle Scientific, UK) in a FastPrep™-24 system (MP Biomedicals, OH) at 6.0 m/s for 40 seconds (two cycles, 5-minute cooling period). The homogenized solution was centrifuged (14,000 rpm, 2 min), and the supernatant treated with 1 µL RNase Cocktail™ (Thermo Fisher Scientific) at 65°C for 15 minutes, then cooled on ice (15 min). After adding 20 µL Proteinase K, samples were vortexed with Qiagen AL lysis buffer (Qiagen, Netherlands) and incubated at 56°C for 10 minutes. DNA was precipitated with 96–100% ethanol, purified using Qiagen spin columns, and eluted in 40 µL Elution Buffer. DNA concentration was measured via Qubit 2.0 fluorometer (Thermo Fisher Scientific). Library preparation followed LITE and LITE 2.0 protocols, and WGS was performed on the Illumina NovaSeq platform, generating 150 base pair (bp) paired end reads. Samples with <30× coverage were excluded to ensure data quality. Prospective *A. fumigatus* WGS data were consolidated with an existing UK WGS collection (616 genomes).

### Sequence alignment and variant calling

Raw 150 bp paired-end reads were quality-checked using FastQC (Babraham Institute, v0.10.1) and aligned to the *A. fumigatus* AF293 reference genome ^83^ using BWA-MEM (v0.7.15) ^84^. SAMtools was used for BAM conversion, sorting and indexing, while Picard Tools (v2.6.0) was used to assign read group information and mark duplicate reads for exclusion. Variant calling was performed with GATK HaplotypeCaller (v4.0) using a ploidy setting of 1, excluding repetitive regions identified by RepeatMasker ^85^ via the ‘-XL’ parameter. Variants were filtered with a minimum confidence threshold of 30 and base quality of 20, and Base Quality Score Recalibration was applied. High-confidence SNPs were retained using GATK VariantFiltration with the following thresholds: QD < 2.0, FS > 60.0, MQ < 40.0, MQRankSum < - 12.5, ReadPosRankSum < -8.0, and SOR > 4.0. SNPs failing these criteria were removed. Non-synonymous SNPs (nsSNPs) were annotated using VCFannotator (Broad Institute). After quality filtering, clinical and environmental WGS data were integrated into a UK-wide genomic repository, expanding the dataset to 912 genomes. Genomes were compared using genome-wide SNP analysis and phylogenetics to identify closely related isolate pairs and infer potential transmission pathways

### Genomic diversity analysis

Phylogenetic trees were inferred from whole genome SNP alignments using RAxML-NG ^86^ with the GTR+G substitution model. Bootstrap replicates were concatenated until convergence was reached, determined by the by the --bsconverge command. Trees were visualised in R v4.3.1 *via* ggTree ^87^ and ggTreeExtra ^88^. Multivariate analyses were performed in R v4.3.1 using adegenet (v2.1.10) following established protocols ^89–92^. FASTA alignments were converted into a genind object, applying a polymorphism threshold of 0.01 to account for low frequency variants. Population groups were assigned, and missing data were imputed using mean allele frequencies *via* the NA.method=’mean’ function. Discriminate analysis of principle components (DAPC) based on SNPs was conducted to assess genetic differentiation among isolates. The optimal number of principal components (PCs) was selected using cross-validation (‘xvalDapc’), based on the lowest mean squared error (MSE). DAPC results were visualized as scatter plots, with population groups mapped to data points and an inset plot showing the cumulative variance explained by PCs. Reference genomes were included as controls.

## Data Availability

All data produced in the present study are available upon reasonable request to the authors

https://microreact.org/project/ohanRa45Se2kRQQugq6mK9-uk

## Ethics

The London–Brent Research Ethics Committee of the United Kingdom Health Research Authority gave ethical approval for this work (REC reference: 19/LO/1663).

