## Supplementary figures and images for "Genomic epidemiology links azole-resistant *Aspergillus fumigatus* hospital bioaerosols to chronic respiratory aspergillosis"

### Supplemental Figure 1

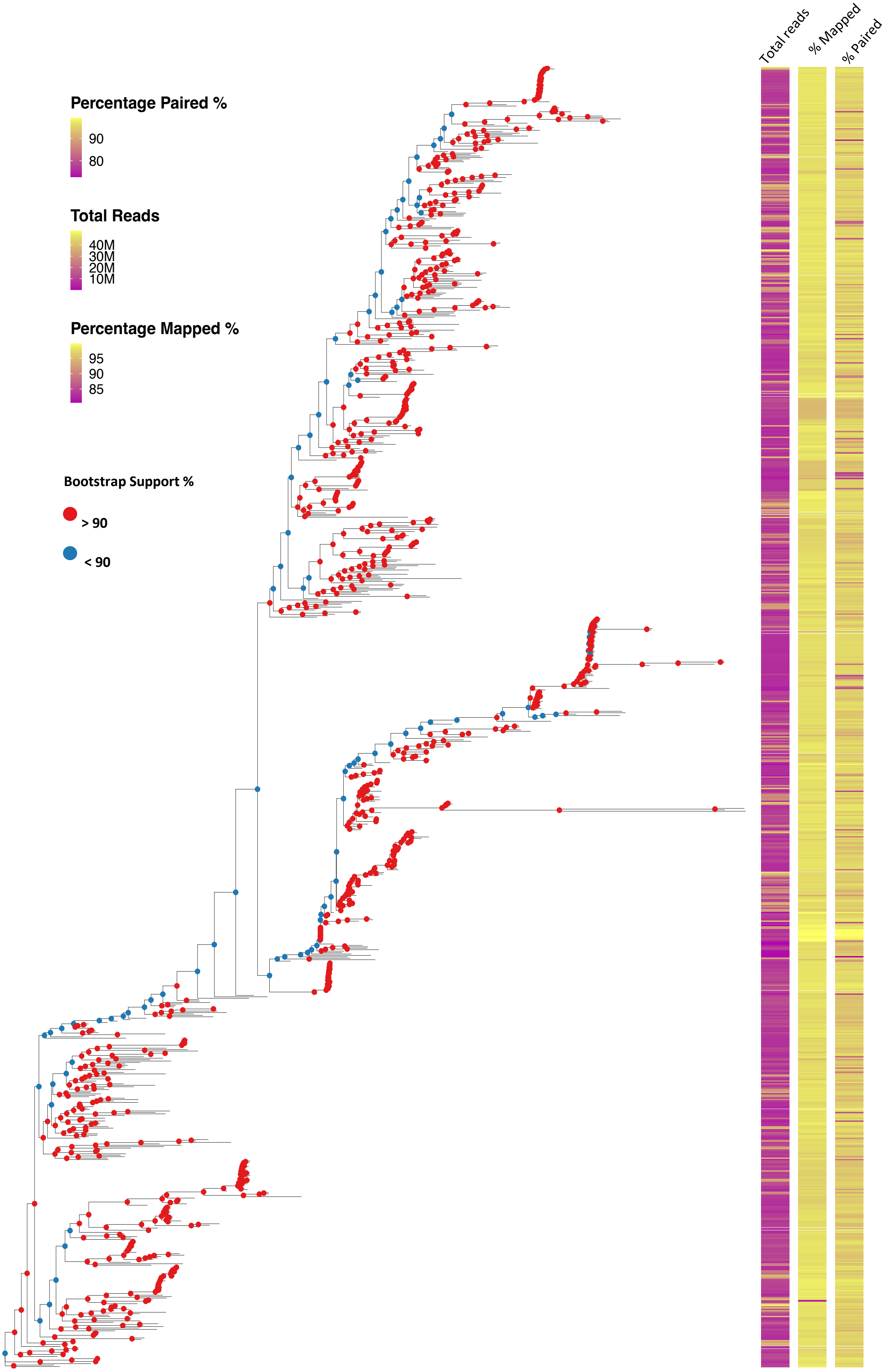
