## Supplemental Table 1 for "Genomic epidemiology links azole-resistant *Aspergillus fumigatus* hospital bioaerosols to chronic respiratory aspergillosis"

| Case ID | Diagnosis | m/f | Antifungal treatment history |
|---|---|---|---|
| Case 30 | CF / CPA | f | Voriconazole, Posaconazole, Isavuconazole and Caspofungin |
| Case 31 | CPA | f | Isavuconazole, Micafungin |
| Case 32 | CPA | f | Caspofungin |
| Case 33 | CPA | f | Itraconazole, Posaconazole, Caspofungin |
| Case 34 | CPA | f | Itraconazole, Voriconazole, Posaconazole, Isavuconazole, caspofungin |
| Case 35 | CPA | f | Itraconazole, Posaconazole, Isavuconazole, Caspofungin |
| Case 36 | ABPA | m | Voriconazole |
| Case 37 | CF | f | Posaconazole |
| Case 38 | CPA | m | Itraconazole |
| Case 39 | CF | m | Itraconazole, Voriconazole, Posaconazole, Isavuconazole, Caspofungin |
| Case 40 | CF | m | Itraconazole |
| Case 41 | ABPA | f | Itraconazole, Posaconazole, Caspofungin |
| Case 42 | ABPA | f | Itraconazole, Posaconazole |
| Case 43 | ABPA | m | Itraconazole, Voriconazole, Posaconazole |
| Case 44 | *Af.* Colonisation | f | nil |
| Case 60 | CPA | m | Itraconazole, Posaconazole |
| Case 61 | ABPA | m | Itraconazole |
| Case 45 | CPA | m | Itraconazole, Voriconazole |
| Case 46 | CPA | f | Itraconazole, Posaconazole |
| Case 47 | ABPA | f | nil |
| Case 48 | CPA | f | Itraconazole |
| Case 49 | ABPA | m | Itraconazole |
| Case 50 | ABPA | f | nil |
| Case 51 | COPD | m | nil |
| Case 52 | ABPA | f | Itraconazole, Voriconazole |
| Case 53 | ABPA | m | nil |
| Case 54 | CPA | m | Itraconazole, Posaconazole |
| Case 55 | CF | f | Posaconazole |
| Case 56 | ABPA | f | nil |
| Case 57 | CF | m | nil |
| Case 58 | CF | f | nil |
| Case 59 | CPA | f | Posaconazole |
| Case 60 | ABPA | m | Itraconazole |
| Case 61 | CPA | f | Posaconazole, Caspofungin |
| Case 62 | CPA | m | Voriconazole, Posaconazole, Isavuconazole and Caspofungin |
| Case 63 | ABPA | f | nil |
| Case 64 | CF | f | Itraconazole |
| Case 65 | CPA | f | Itraconazole, Posaconazole, Caspofungin |
| Case 66 | CF | f | Itraconazole, amphotericin nebulised therapy |
| Case 67 | CPA | f | Itraconazole, Posaconazole, Caspofungin |
| Case 68 | CPA | m | Itraconazole, Voriconazole |
| Case 69 | ABPA | f | Itraconazole, Posaconazole |
| Case 70 | CPA | f | nil |
| Case 71 | ABPA | f | Itraconazole, Voriconazole, Posaconazole |
| Case 72 | ABPA | f | nil |
| Case 73 | ABPA | f | Itraconazole, Posaconazole |
| Case 74 | CPA | m | Itraconazole, Voriconazole, Posaconazole, Caspofungin |
| Case 75 | CPA | m | Voriconazole |
| Case 76 | CPA | m | Itraconazole, Posaconazole, Isavuconazole, Caspofungin |
| Case 77 | ABPA | f | nil |
| Case 78 | ABPA | m | Itraconazole |
| Case 79 | ABPA | f | Itraconazole, Posaconazole |
| Case 80 | ABPA | m | Itraconazole |
| Case 81 | ABPA | m | Itraconazole |
| Case 82 | ABPA | f | Itraconazole |
